# Regional Service-System Conditions Associated with Facility-Linked Home-Based Specialist Care in Japan: A Claims-Based Ecological Study of Home Dialysis

**DOI:** 10.64898/2026.06.17.26355849

**Authors:** Kenta T. Suzuki

## Abstract

**Background:** Complex chronic care is increasingly delivered in patients’ homes while remaining linked to specialist facilities for training, monitoring, and backup care. Home dialysis provides a useful case because peritoneal dialysis (PD) and home hemodialysis (HHD) share a home–facility delivery structure but differ in technical and operational requirements. This study examined regional service-system conditions associated with the presence and scale of PD and HHD in Japan.

**Methods:** This ecological study used publicly available claims, administrative, census, and geospatial data harmonized to 334 Secondary Medical Areas. Regional indicators were organized into four domains: dialysis service delivery, implementation support for home-based care, hospital backup capacity, and living and sociodemographic context. Diffusion was examined using claims-based indicators of regional presence and post-presence scale, analyzed separately for PD and HHD with Firth penalized logistic regression and zero-truncated negative binomial regression, respectively.

**Results:** PD was observed in 271 regions and HHD in 109. Patterns of associated regional conditions differed by modality and stage. PD was associated mainly with existing dialysis-service organization, whereas HHD was associated with broader regional supports, including home-care delivery, living infrastructure, transition support, and hospital-system indicators. Conditions associated with presence differed from those associated with scale. Cross-modality associations suggested that shared regional factors may shape the distribution of both modalities.

**Conclusions:** Regional conditions for home dialysis diffusion in Japan differed by modality and stage. PD was linked mainly to existing dialysis-service organization, whereas HHD was linked to multi-domain regional support for technically demanding home treatment. Under standardized reimbursement, local service-system capacity may remain important for modality- and stage-specific diffusion of home dialysis.

## INTRODUCTION

Chronic care increasingly includes arrangements that span the home and specialist facilities [1, 2]. These arrangements separate the site where treatment is performed from the systems that sustain treatment delivery. This structure is especially visible in technically demanding treatments such as home mechanical ventilation, outpatient parenteral antimicrobial therapy, and home dialysis [3–6]. In these treatments, key activities occur in the home, while supporting clinical services organize training, supplies, monitoring, technical support, and escalation [3, 4, 6]. These supports must be sustained over time and across patients [3, 4, 6, 7]. Regional diffusion of such treatments therefore depends not only on treatment characteristics and clinical need, but also on the capacity of local service systems to sustain safe delivery across home and specialist-facility settings [8–10].

Home dialysis provides a useful empirical case for examining this service-system dependence within a single treatment field. Its two principal modalities, peritoneal dialysis (PD) and home hemodialysis (HHD), share a home–facility delivery structure but differ in operational demands. PD is typically organized around self-care routines, supply management, and periodic facility oversight [11, 12]. HHD requires more intensive training, equipment installation, water and drainage arrangements, and reliable technical and escalation support [4, 13, 14]. These HHD requirements have been identified as barriers to uptake [7, 15, 16], and HHD has diffused less widely than PD in many health systems [7, 17–20]. These differences suggest that PD and HHD may depend on partly different regional service-system conditions, despite their shared home–facility delivery structure.

Prior work has identified multiple influences on home dialysis use, including local resources, health-system policy, provider factors, lifestyle considerations, and care-partner burden [7]. Empirical studies have also examined geographic, facility-, and center-level variation in home dialysis delivery and use [21–23]. However, these factors have less often been examined together as multidomain regional service-system conditions within a common framework. It therefore remains unclear whether the operational differences between PD and HHD correspond to different regional service-system conditions, or whether such conditions differ between initial regional presence and subsequent scale of use.

Two questions follow from this analytic gap. First, do the regional service-system conditions associated with regional presence differ from those associated with post-presence scale? Second, do the regional conditions associated with HHD diffusion resemble those associated with PD, or do they reflect HHD’s higher operational demands?

Japan provides a useful setting for examining regional service-system conditions because insured care is paid under a national fee schedule, limiting regional variation in the basic payment framework [24–26]. This makes payment-system variation a less likely competing explanation for regional differences in diffusion. Secondary Medical Areas are statutory planning units for regional inpatient care and provide a policy-relevant frame for examining regional variation in service conditions [27]. Because home dialysis depends on facility oversight and hospital backup [4, 7, 13], this frame is relevant to the regional systems that support PD and HHD diffusion.

This study examined home dialysis diffusion in Japan as a regional service-system phenomenon using Secondary Medical Areas as the analytic unit. Regional indicators were organized into four domains: dialysis service delivery, implementation support for home-based care, hospital backup capacity, and living and sociodemographic context. Standardized national health-insurance claims data were used to measure regional presence and post-presence scale for PD and HHD. The analysis assessed whether the patterns of regional conditions associated with diffusion were shared across modalities and stages, or were instead consistent with modality-specific requirements, stage-specific processes, or a more general regional capacity for facility-linked home-based specialist care.

## MATERIALS and METHODS

### Data Source

Indicators were constructed from publicly available datasets. Sociodemographic and housing data came from the 2020 Population Census of the Statistics Bureau of Japan. Medical resource data came from the Ministry of Health, Labour and Welfare, including the Survey of Medical Institutions [28] and the NDB Open Data [29]. The NDB is Japan’s national database of health insurance claims and specific health checkups. Facility standard notifications were obtained from the Regional Bureaus of Health and Welfare. Geographic data came from the Geospatial Information Authority of Japan.

Outcome variables and covariates were aligned to Japan’s fiscal year (FY) 2023. When FY2023 data were unavailable, the most recent available year was used. The earliest source was the 2020 Population Census. Detailed definitions, calculation formulas, sources, and reference years for all variables are listed in **Supplementary Table 1**. All datasets were anonymized and publicly accessible. Institutional review board approval and informed consent were therefore not required.

### Study unit and data harmonization

Japan’s Secondary Medical Areas served as the analytic units. Of the 335 SMAs nationwide, Kawasaki North and Kawasaki South were combined because of data constraints, yielding 334 analytic units.

Data reported at facility, municipality, and SMA levels were harmonized to the SMA level. Municipality-level data were aggregated to SMAs using an official municipality–SMA correspondence table. Identification, record linkage, and geocoding procedures for dialysis facilities and home-visit nursing agencies are described in the **Supplementary Methods**.

### Definitions of hemodialysis and home-based dialysis

Hemodialysis (HD), home-based dialysis, and regional home-care systems were defined using reimbursement claim items from the 10th NDB Open Data. The primary indicators were the number of claims for each item and, where available, the number of patients, defined as the annual unduplicated count of individuals with at least one claim for the item during the fiscal year.

Home-based dialysis was classified into PD and HHD. PD was defined by the number of patients billed for the home peritoneal dialysis self-care instruction and management fee. Because this indicator includes both continuous ambulatory and automated peritoneal dialysis and may include patients receiving concurrent facility-based HD, it was treated as an approximate measure of PD use including combination therapy, rather than as a direct estimate of exclusive PD volume. HHD was defined by the number of patients billed for the home hemodialysis instruction and management fee.

Regional HD volume was estimated using a previously published formula [30]. Because this estimate may structurally include patients on home hemodialysis, it was treated as an approximate measure of overall hemodialysis volume rather than a precise count of facility-based HD patients. Details of the modification and implementation are provided in the **Supplementary Methods**.

Indicators of regional home-care systems were constructed from the comprehensive home medical management fee and the discharge joint guidance fee (Type 2), which primarily reflect comprehensive management of home-care patients and support for hospital-to-home transitions. These items served as proxy measures of the breadth and intensity of regional home-care delivery. Names and claim codes of all reimbursement items are listed in **Supplementary Table 2**.

### Statistical Analysis

#### Covariates and variable construction

Explanatory variables were organized into four conceptual domains: the dialysis service delivery system, implementation support for home-based care, hospital backup capacity, and the living and sociodemographic context. Each domain comprised indicators intended to capture the corresponding aspect of regional system capacity. The indicators within and across these domains spanned different levels of dialysis specificity, ranging from dialysis-specific claims measures to general regional context measures, and this heterogeneity was retained intentionally to capture the multidomain character of regional service-system capacity. Complete variable definitions and specificity levels are provided in **Supplementary Table 3**.

For the adoption models, explanatory variables were expressed as population-standardized measures or proportions and were then standardized to zero mean and unit variance; adjusted odds ratios therefore correspond to a one-standard-deviation increase in each covariate unless otherwise noted. For the scale models, the logarithm of the total dialysis patient count was included as an offset, and explanatory variables were entered on their original scales as counts or proportions to avoid double adjustment for population size; incidence rate ratios correspond to one-unit increases as coded.

To reduce simultaneity, PD and HHD claim counts from FY2022 were entered as cross-modality predictors only: prior PD volume into HHD models, and prior HHD volume into PD models. Same-modality lags were not used, as they would predict the current outcome from a near-copy of itself. This specification tests whether regional experience with one home modality relates to uptake of the other. Other variables were aligned to FY2023, or to the most recent available year.

Multicollinearity among candidate variables was assessed using Pearson correlation coefficients. For variable pairs with strong correlations (|r| > 0.7) [31], one variable was selected with priority given to conceptual relevance and interpretability. Variance inflation factors (VIF) were subsequently examined in the final models, and further variable removal was performed when the variance inflation factor exceeded 7. Collinearity screening and VIF-based removal were performed separately within each of the four models; consequently, the final covariate sets differ across models.

#### Two-part model for adoption and scale

Regional variation in home-based dialysis was analyzed in two stages [10]: claims-based regional adoption and post-adoption scale. Adoption was defined as recorded regional presence based on a non-suppressed positive claim count for the corresponding home-dialysis management fee. It therefore represents regional presence rather than incident initiation of PD or HHD. Scale was defined as the post-adoption intensity of claims among adopting SMAs. It represents claims-based use after adoption rather than registry-equivalent modality prevalence. This distinction allows regional conditions associated with the presence of a modality to be distinguished from those associated with its level of claims-based use after adoption. The two-part framework was applied separately to PD and HHD, yielding four models in total.

For adoption, the outcome was a binary indicator of whether a non-suppressed positive claim count for PD or HHD was observed in the public NDB aggregate data for a given SMA during FY2023. Adoption was modeled using Firth penalized logistic regression [32], which reduces bias in maximum likelihood estimation and addresses potential separation in multivariable logistic models. Because the PD and HHD management fees are generally billable once per patient per month, the NDB suppression threshold of ten claims per year implies that a non-suppressed positive claim count corresponds to at least 11 patient-months of billing activity in the region. The claims-based adoption indicator should therefore be interpreted as sustained regional billing activity for home dialysis, rather than as a measure of incident initiation or a stringent volume threshold [29].

For scale, the outcome was the annual number of claims for PD or HHD among adopting SMAs. Scale was modeled using zero-truncated negative binomial regression [33], with the logarithm of the total number of dialysis patients included as an offset to account for regional differences in dialysis volume. Under this specification, regression coefficients reflect the intensity of home-based dialysis claims per dialysis patient. Given the monthly billing structure of the PD and HHD management fees, this outcome should be interpreted as an indicator of home-dialysis patient-month intensity rather than as a direct estimate of patient share.

Model adequacy was assessed using residual diagnostics and goodness-of-fit measures. Moran’s I statistic [34] was calculated for model residuals to examine spatial autocorrelation, with spatial weights defined using queen contiguity and row-standardized.

#### Sensitivity analyses

Sensitivity analyses assessed robustness to two analytic choices. First, proximity-based indicators were recalculated using alternative distance thresholds of 3 km and 10 km, in addition to the main 5-km threshold. These analyses were applied to the adoption and scale models while keeping all other model specifications unchanged.

Second, because PD adoption was observed in most SMAs under the claims-based definition, the PD adoption model was re-estimated using a patient-based definition of presence. Under this definition, SMAs were classified as adopting PD when a non-suppressed patient count for the home peritoneal dialysis self-care instruction/management fee was reported in FY2023. Because public NDB Open Data suppresses small counts, this definition was interpreted as a more stringent patient-based presence indicator rather than as a simple claim-presence threshold.

#### Software and significance

Analyses were conducted in R 4.5.1 and Python 3.12. The random seed was set to 2026. Firth penalized logistic regression, zero-truncated negative binomial regression, Moran’s I and LISA statistics [34, 35], and interval regression were implemented in R using the logistf, pscl, spdep, and survival packages, respectively. Data harmonization, geocoding, and spatial joins were performed in Python using pandas and geopandas. Effect estimates are reported with 95% confidence intervals. Two-sided p-values below 0.05 were considered statistically significant.

Inference focused on cross-model and cross-domain patterns rather than on individual coefficients, as covariates within each domain were intended to capture overlapping aspects of regional capacity rather than to function as independent candidates. No formal correction for multiple comparisons was therefore applied to the primary analysis [36]. Benjamini–Hochberg [37] q values were computed within each model and are reported in the Supplementary Tables.

## RESULTS

### Regional Characteristics by Dialysis Modality Availability

**Table 1** summarizes SMA characteristics according to claims-based adoption of home dialysis in FY2023. Adoption was defined as the presence of at least one claim for the corresponding reimbursement item during FY2023. PD was observed in 271 of 334 SMAs (81.1%) and HHD in 109 (32.6%), indicating broader recorded regional presence of PD and more limited diffusion of HHD. Descriptively, SMAs with recorded PD or HHD use tended to be more urbanized than those without recorded use, as reflected in higher DID share and lower aging rate, with this contrast particularly pronounced for HHD. HHD-observed SMAs also showed higher home-care management volume and sewerage coverage.

**Table 1.** Regional characteristics by PD/HHD availability. Values are presented as median (interquartile range) unless otherwise indicated. PD and HHD availability was defined as whether at least one corresponding claim was observed during FY2023. Covariates were selected according to the criteria described in the Methods; descriptive statistics and regression analyses were conducted using the retained variables. The correlation matrix for all candidate covariates is shown in **Supplementary Figure 1**. “–” indicates not applicable because PD-and HHD-related variables are shown only within strata of the corresponding modality. Abbreviations: CE, clinical engineer; HD, hemodialysis; HHD, home hemodialysis; HVN, home-visit nursing agency; PD, peritoneal dialysis.

|  | Overall<br>N = 334<sup>1</sup> | PD observed |  | HHD observed |  |
| --- | --- | --- | --- | --- | --- |
|  |  | No<br>N = 63<sup>1</sup> | Yes<br>N = 271<sup>1</sup> | No<br>N = 225<sup>1</sup> | Yes<br>N = 109<sup>1</sup> |
| Dialysis patients | 629<br>(297, 1,359) | 224<br>(150, 365) | 804<br>(387, 1,569) | 440<br>(233, 855) | 1,447<br>(735, 2,267) |
| PD volume | 134<br>(31, 450) | - | 228<br>(79, 537) | 78<br>(12, 236) | 425<br>(198, 1,075) |
| HHD volume | 0<br>(0, 12) | - | 0<br>(0, 24) | - | 36<br>(12, 97) |
| Dialysis facility density | 4.77<br>(3.82, 5.95) | 5.62<br>(4.15, 6.76) | 4.61<br>(3.74, 5.70) | 4.93<br>(3.87, 6.31) | 4.54<br>(3.78, 5.43) |
| CEs density | 20<br>(15, 26) | 16<br>(9, 24) | 20<br>(16, 26) | 19<br>(14, 26) | 22<br>(17, 26) |
| Weekend HD share | 12.2<br>(8.8, 15.9) | 7.9<br>(6.9, 12.2) | 13.1<br>(9.8, 16.6) | 11.1<br>(7.6, 14.7) | 14.0<br>(12.0, 17.1) |
| Home care management volume | 406<br>(240, 597) | 308<br>(152, 471) | 419<br>(271, 668) | 360<br>(198, 516) | 562<br>(389, 779) |
| Nearest HVN distance | 426<br>(262, 690) | 616<br>(160, 1,015) | 407<br>(264, 633) | 495<br>(288, 769) | 341<br>(242, 495) |
| Discharge coordination rate | 1.20<br>(0.23, 2.02) | 0.00<br>(0.00, 1.82) | 1.31<br>(0.64, 2.12) | 1.11<br>(0.00, 1.90) | 1.51<br>(0.95, 2.21) |
| HVN availability | 9<br>(4, 24) | 4<br>(2, 7) | 12<br>(5, 28) | 7<br>(3, 13) | 23<br>(11, 53) |
| HVN density | 11.6<br>(9.0, 15.2) | 11.0<br>(8.1, 14.5) | 11.6<br>(9.2, 15.3) | 11.1<br>(8.7, 14.8) | 12.2<br>(10.2, 15.3) |
| General hospital bed density | 718<br>(587, 882) | 720<br>(605, 832) | 718<br>(581, 892) | 720<br>(587, 882) | 717<br>(603, 876) |
| Regional tertiary hospital density | 0.55<br>(0.24, 0.86) | 0.00<br>(0.00, 0.87) | 0.57<br>(0.37, 0.86) | 0.51<br>(0.00, 0.88) | 0.57<br>(0.43, 0.85) |
| Habitable population density | 650<br>(336, 1,330) | 330<br>(188, 503) | 771<br>(402, 1,606) | 457<br>(288, 843) | 1,336<br>(664, 4,241) |
| DID share | 40<br>(16, 65) | 15<br>(0, 30) | 47<br>(23, 72) | 29<br>(13, 49) | 65<br>(43, 91) |
| Taxable income per taxpayer | 3,283<br>(3,059, 3,584) | 3,120<br>(2,927, 3,283) | 3,328<br>(3,081, 3,673) | 3,199<br>(2,999, 3,391) | 3,559<br>(3,268, 3,937) |
| Homeownership rate | 69<br>(63, 76) | 76<br>(70, 79) | 68<br>(61, 74) | 71<br>(66, 77) | 65<br>(59, 71) |
| Single-person household share | 33.2<br>(29.8, 37.1) | 32.2<br>(28.5, 37.1) | 33.4<br>(30.0, 37.1) | 32.4<br>(29.1, 35.9) | 34.8<br>(30.9, 38.5) |
| Population aged ≥65 years | 31.7<br>(28.3, 35.4) | 36.6<br>(32.2, 39.2) | 30.8<br>(27.7, 34.1) | 33.5<br>(29.9, 37.0) | 28.9<br>(26.3, 31.6) |
| LTC certification rate | 4.50<br>(3.54, 5.57) | 4.20<br>(3.15, 5.41) | 4.59<br>(3.67, 5.60) | 4.27<br>(3.38, 5.18) | 5.33<br>(3.95, 6.10) |
| Sewerage coverage | 69<br>(44, 85) | 50<br>(27, 69) | 72<br>(51, 89) | 57<br>(40, 78) | 83<br>(69, 97) |

<sup>1</sup> Median (Q1, Q3); n (%)

Spatial patterns in the PD- and HHD-to-HD claim ratios differed between modalities (**Figure 1**). PD showed no significant global spatial autocorrelation (Moran’s I = −0.0242, p = 0.713), whereas HHD showed weak but significant positive autocorrelation (Moran’s I = 0.1025, p = 0.002). Local indicators of spatial association provided little evidence of substantial clustering for PD and only limited local clustering for HHD (**Supplementary Figures S1 and S2**). PD was broadly distributed across SMAs, whereas HHD showed modest geographic clustering.

**Figure 1.**
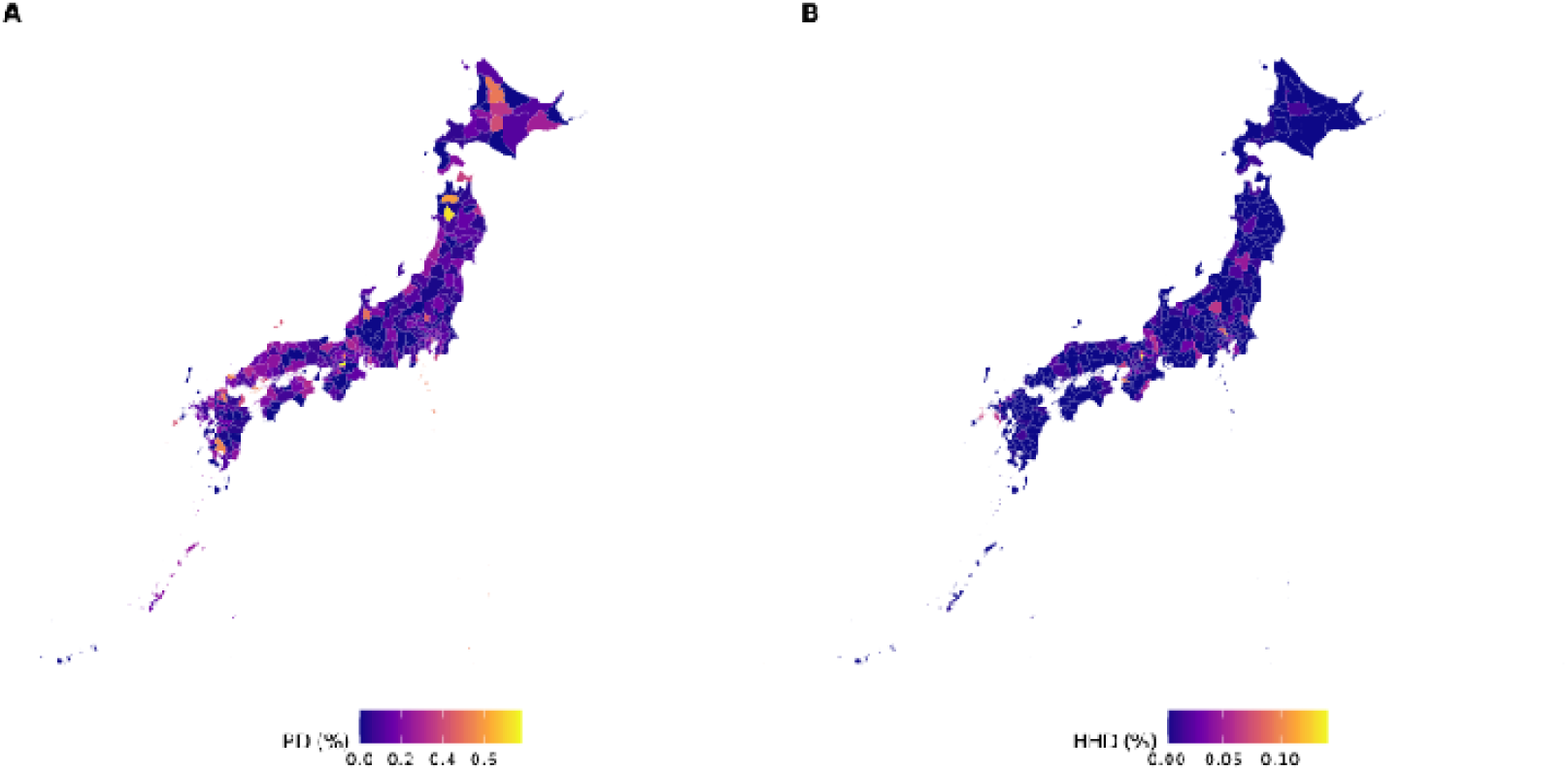
Regional distribution of the ratios of PD and HHD claims to hemodialysis claims across Secondary Medical Areas in Japan. Panel A shows the ratio of PD claims to hemodialysis claims, and Panel B the ratio of HHD claims to hemodialysis claims, for each SMA. In both panels, the numerator was the number of claims for the corresponding home dialysis reimbursement item and the denominator was the number of hemodialysis claims in the same SMA. Because hemodialysis claims are billed repeatedly whereas the relevant home dialysis management fees are generally billed once monthly, these ratios should be interpreted as claims-based relative intensity measures rather than as direct estimates of patient share. Maps are shown at the SMA level.

### Two-stage analysis of adoption and scale

The two-part framework was used to distinguish factors associated with whether a modality was present in a region from those associated with the intensity of use after adoption. Results are therefore presented separately by stage and modality. Only variables retained in the final multivariable models are reported here; the full candidate set and screening procedure are described in the Methods and **Supplementary Table 3**.

In the adoption-stage models (**Figure 2**), the factors associated with regional presence differed by modality. In the PD adoption model, Weekend HD share (adjusted OR 1.13, 95% CI 1.04–1.24) and HVN availability (adjusted OR 2.75, 95% CI 1.57–4.96) were positively associated with the presence of PD claims. In contrast, in the HHD adoption model, Lagged PD volume (adjusted OR 1.73, 95% CI 1.14–2.74), Home care management volume (adjusted OR 1.56, 95% CI 1.03–2.37), and Sewerage coverage (adjusted OR 1.02, 95% CI 1.00–1.04) were positively associated with adoption. Thus, the correlates of adoption showed limited overlap between PD and HHD.

**Figure 2.**
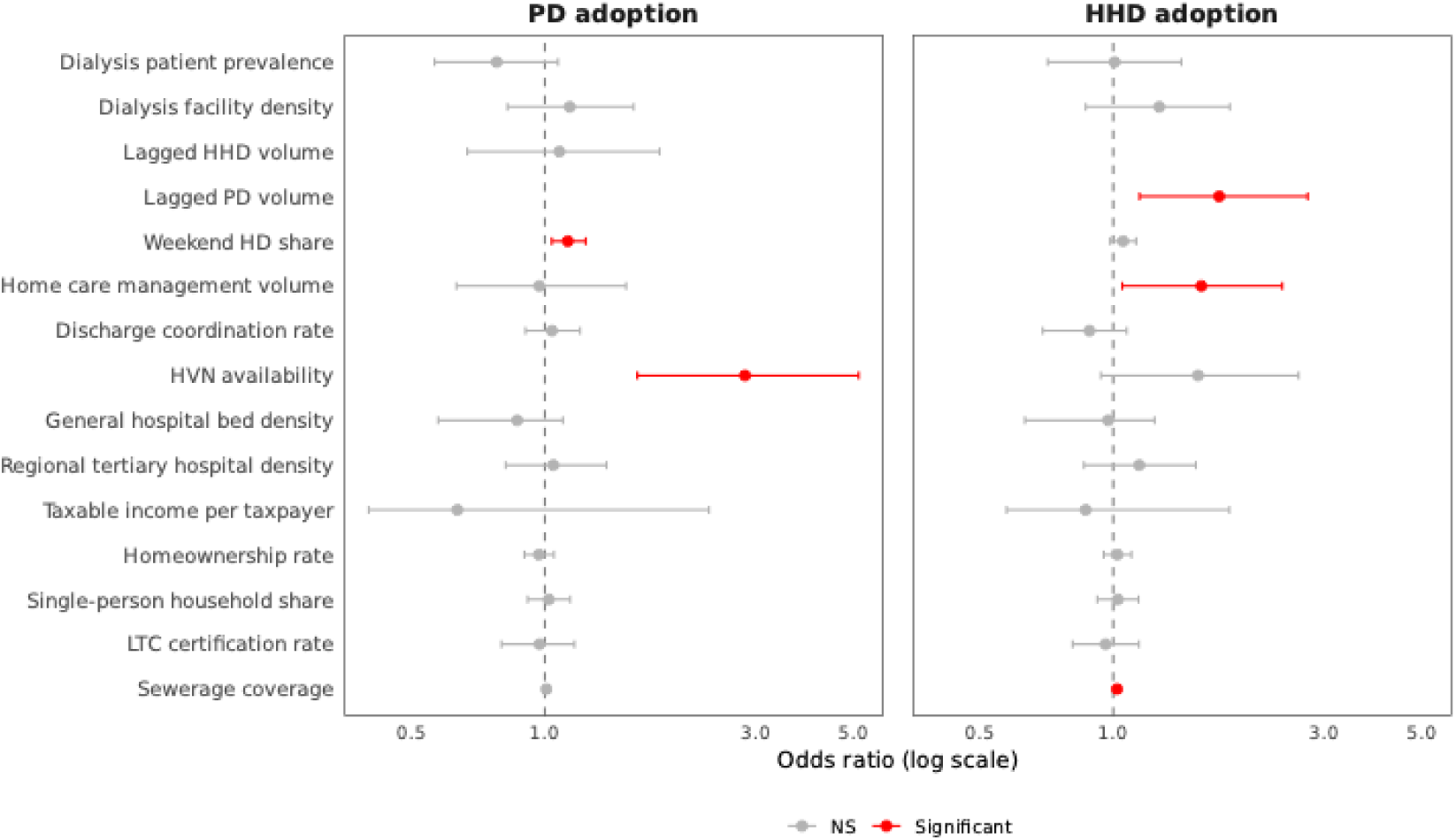
Factors associated with adoption of PD and HHD across SMAs. Forest plots show adjusted odds ratios and 95% confidence intervals from multivariable Firth penalized logistic regression models for modality adoption. The left panel shows PD adoption and the right panel HHD adoption. Values above 1 indicate a higher likelihood that the modality was present in an SMA, and values below 1 indicate a lower likelihood. Colored points indicate associations with p < 0.05; gray points indicate associations not meeting this threshold. Full model results are provided in **Supplementary Tables 4 and 5**.

Among adopting SMAs, the correlates of scale differed by modality and did not simply reproduce the adoption-stage patterns **(Figure 3**). For PD, Dialysis facility density was negatively associated with scale (adjusted IRR 0.67, 95% CI 0.53–0.84), whereas Lagged HHD volume (adjusted IRR 1.14, 95% CI 1.01–1.29) and Single-person household share (adjusted IRR 1.03, 95% CI 1.01–1.05) were positively associated. For HHD, Discharge coordination rate was positively associated with scale (adjusted IRR 1.33, 95% CI 1.11–1.60), whereas Regional tertiary hospital density was negatively associated (adjusted IRR 0.64, 95% CI 0.47–0.87). These patterns did not simply reproduce those observed for adoption. Full multivariable results, including FDR-adjusted q values for individual coefficients, are provided in **Supplementary Tables 4–7**.

**Figure 3.**
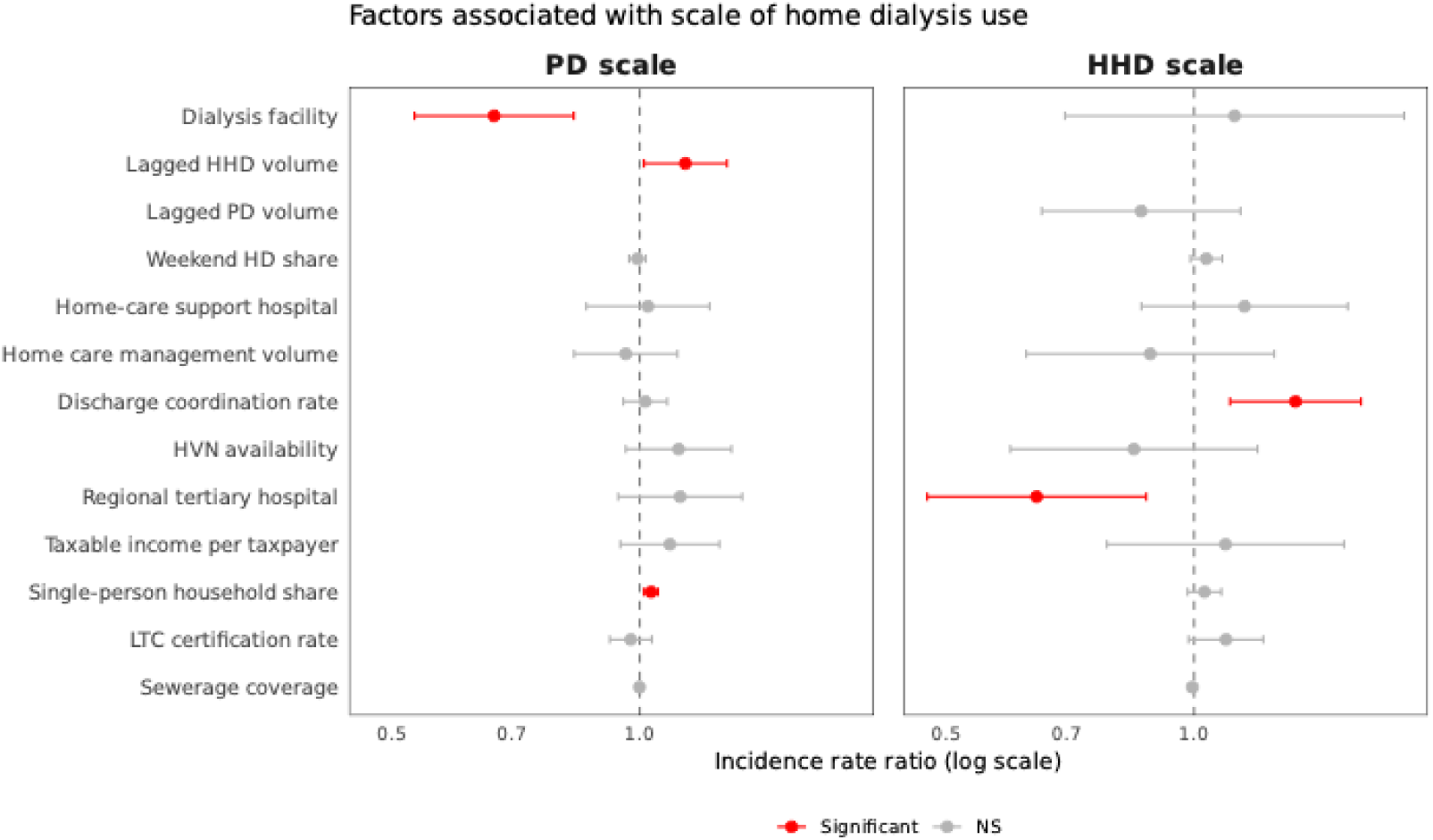
Factors associated with the scale of PD and HHD use among adopting SMAs. Forest plots show adjusted incidence rate ratios and 95% confidence intervals from zero-truncated negative binomial models estimated among SMAs in which the modality was present. The left panel shows PD scale and the right panel HHD scale. Values below 1 indicate lower claim intensity, and values above 1 indicate higher claim intensity. Filled points indicate associations with p < 0.05; unfilled points indicate associations not meeting this threshold. Full model results are provided in **Supplementary Tables 6and 7**.

Taken together, these findings indicate that the patterns of regional conditions associated with home dialysis diffusion differed by stage and modality. The factors associated with whether a modality was present in an SMA were not the same as those associated with intensity of use after adoption, and these patterns also differed between PD and HHD. These cross-model contrasts are summarized in **Table 2**.

**Table 2.** Summary of factors associated with home dialysis diffusion across stages and modalities. This table summarizes model-specific associations retained in the final multivariable models and is intended to highlight cross-model patterns across modality and diffusion stage. Adoption of peritoneal dialysis (PD) and home hemodialysis (HHD) across Secondary Medical Areas in Japan was analyzed using Firth penalized logistic regression, with estimates reported as adjusted odds ratios (ORs) and 95% confidence intervals. Scale among adopting SMAs was analyzed using zero-truncated negative binomial regression, with estimates reported as adjusted incidence rate ratios (IRRs) and 95% confidence intervals. Positive and negative directions indicate whether higher values of each regional factor were associated with greater or lower claims-based modality presence or post-adoption claim intensity, respectively. Note: ORs are per one-SD increase; IRRs are per one-unit increase as coded. Effect magnitudes are therefore not directly comparable across stages; comparisons in the text are directional.

| Stage | Modality | Factor retained in final model | Direction | Adjusted effect size<br>(95 % CI) |
| --- | --- | --- | --- | --- |
| Adoption | PD | Weekend HD share | Positive | OR 1.13 (1.04–1.24) |
| Adoption | PD | HVN availability | Positive | OR 2.75 (1.57–4.96) |
| Adoption | HHD | Lagged PD volume | Positive | OR 1.73 (1.14–2.74) |
| Adoption | HHD | Home care management volume | Positive | OR 1.56 (1.03–2.37) |
| Adoption | HHD | Sewerage coverage | Positive | OR 1.02 (1.00–1.04) |
| Scale | PD | Dialysis facility density | Negative | IRR 0.67 (0.53–0.83) |
| Scale | PD | Lagged HHD volume | Positive | IRR 1.14 (1.01–1.28) |
| Scale | PD | Single-person household share | Positive | IRR 1.03 (1.01–1.05) |
| Scale | HHD | Discharge coordination rate | Positive | IRR 1.33 (1.11–1.60) |
| Scale | HHD | Regional tertiary hospital density | Negative | IRR 0.64 (0.47–0.87) |

### Model diagnostics and sensitivity analyses

Sensitivity analyses showed that the main comparative patterns were generally consistent across alternative outcome definitions and distance thresholds. When PD adoption was redefined using patient-based presence rather than claims-based presence, the overall pattern of associations was broadly similar to the main analysis, and no significant residual spatial autocorrelation was detected. Recalculating proximity-based indicators using 3-km and 10-km thresholds also generally preserved the main directional patterns. In the adoption models, residual spatial autocorrelation was not evident for HHD, whereas some residual spatial structure remained for PD under both alternative distance specifications. In the scale models, residual spatial autocorrelation was not evident for PD or HHD, although diagnostics for HHD scale should be interpreted cautiously because the contiguity structure among adopting SMAs was sparse. Full sensitivity results are shown in **Supplementary Table 9** and **Supplementary Figures 4 and 5**.

## DISCUSSION

This study found that the service-system correlates of home dialysis diffusion differed by diffusion stage and modality, rather than forming a single regional pattern. Within each modality, adoption and scale involved different conditions; across modalities, HHD-related conditions extended beyond dialysis services, whereas PD-related conditions were concentrated in existing dialysis-service organization. Because these contrasts were observed under Japan’s nationally uniform fee-schedule system, shared coverage and payment rules alone are unlikely to produce uniform regional diffusion of complex home-based care. Instead, the associated service-system profiles appear to be modality-and stage-specific [7, 25, 38].

For PD, the associated regional conditions differed between adoption and scale, but in both stages they were mainly located around existing dialysis-service organization. PD adoption was associated with weekend HD share—the proportion of evening and weekend sessions relative to standard HD, indicating flexible service operations—and with HVN availability near dialysis facilities. This pattern suggests that PD adoption was more closely aligned with flexible facility-HD operations and nearby home-support resources than with broader regional support domains. Among adopting regions, PD claim intensity was lower where dialysis facility density was higher, consistent with prior evidence that home dialysis use is lower in regions with higher facility density, typically urban areas [23, 39]. PD scale was also associated with single-person household share, in line with prior evidence that home environment and home-based support are relevant to home dialysis use [39, 40]. PD claim intensity was further elevated where prior HHD activity had been recorded, consistent with a cross-modality regional pattern. Overall, the PD profile was concentrated around facility-HD operations, nearby home-support resources, and regional living context, rather than extending across a broad set of regional support domains.

In contrast, HHD was associated with regional conditions spanning multiple domains beyond the immediate dialysis-service context. At the adoption stage in particular, HHD adoption was associated with prior PD volume, home care management volume, and sewerage coverage. These associations are consistent with the home–facility separation and greater operational requirements that characterize HHD delivery. The observed pattern may reflect regional support infrastructure relevant to HHD initiation and maintenance [4, 13, 14]. Specifically, prior PD volume—potentially proxying accumulated experience with home dialysis—and home care management volume—potentially reflecting the reach of physician-led ongoing home medical care—can be read as regional indicators of this support infrastructure. The association with sewerage coverage is directionally consistent with prior evidence emphasizing water and drainage requirements for HHD [4]. However, sewerage coverage was strongly correlated with urbanicity indicators including population density, DID share, and single-person household share, and whether the observed association reflects housing-level feasibility or urban-context confounding cannot be distinguished here.

At the scale stage, by contrast, the HHD profile was characterized by transition-support and hospital-system indicators. Discharge coordination rate was positively associated with HHD scale, whereas regional tertiary hospital density was negatively associated. The positive association with discharge coordination rate is consistent with the relevance of organized hospital-to-home transition functions after HHD adoption. The negative association with tertiary hospital density should not be interpreted as evidence that tertiary hospitals inhibit HHD expansion; it may instead reflect unmeasured features of hospital-intensive regional systems or referral patterns. Together, these findings are more consistent with a profile involving transition-support capacity than with hospital resource density alone [41, 42]. Overall, the conditions associated with HHD differed between adoption and scale, but in both stages they extended beyond the immediate dialysis-service context to home-care delivery, living infrastructure, transition support, and hospital-system conditions.

Although PD and HHD differ in their associated regional conditions and technical requirements, temporal cross-modality associations were observed between the two modalities. The PD-scale lag was attenuated under FDR adjustment and should be interpreted cautiously; nevertheless, the directional consistency across modalities suggests that activity in one home-dialysis modality may serve as a marker of regional readiness for the other. Prior research has documented substantial facility-level and center-level variation in home dialysis use [7, 21, 22, 43], and plausible explanations include shared regional infrastructure for home-based care or shared provider orientation toward home dialysis on the supply side, and overlapping patient populations who select home dialysis on the demand side. A stepping-stone interpretation is also possible, whereby experience with the less operationally demanding modality contributes to organizational familiarity with facility-linked home dialysis. The ecological design of this study cannot distinguish these possibilities, but they are consistent with integrated home dialysis perspectives that treat PD and HHD as connected home-based options [44, 45].

These findings extend prior evidence of geographic, facility-level, and center-level variation in home dialysis use under formal public coverage [7, 21, 22, 43] by showing that regional variation involves not only overall uptake, but also modality- and stage-specific combinations of service-system conditions. Because Japan’s basic reimbursement rules for PD and HHD are standardized nationally [25], these contrasts are unlikely to be explained by geographic variation in payment rules alone. Instead, they suggest that regional dialysis-service organization, home-care support, hospital transition functions, and community living context may all contribute to how home dialysis diffuses across regions.

The contrast between PD and HHD may also inform how technically demanding home-based treatments are evaluated beyond dialysis. The broader implication is that regional diffusion should be assessed not only by specialist facility characteristics, but also by the regional functions that sustain home delivery, including coordination between specialist facilities and home-based services. When multiple home-based modalities coexist within a therapeutic field, experience or infrastructure in one modality may also indicate regional readiness for another. Similar considerations may apply to home mechanical ventilation and home-delivered outpatient parenteral antimicrobial therapy, which require training, equipment, monitoring, escalation pathways, and home–hospital coordination [3, 5, 6].

## Limitations

This study has several limitations. First, the cross-sectional ecological design at the SMA level does not support patient-level or causal inference. Observed associations should be interpreted as between-area correlations rather than as individual-level determinants or direct effects of policy intervention. Unmeasured factors—including patient preference, care-partner availability, home environment suitability, modality education, referral practices, facility-level experience, and provider culture—may have contributed to residual confounding, and SMAs are health-planning units rather than closed care markets, with patients potentially receiving dialysis-related care across area boundaries.

Second, outcomes were based on publicly available claims aggregates rather than patient-level modality use. Because cells with ten or fewer claims were suppressed and treated as zero, and the PD and HHD management fees are generally billable once per patient per month, the claims-based adoption indicator corresponds to at least 11 patient-months of regional billing activity. It does not identify the number of patients or distinguish one patient treated for most of the year from multiple patients treated for shorter periods. Misclassification is therefore most likely in regions with low or intermittent billing activity. PD claims may include patients on concurrent facility-based HD and represent claims-based use rather than stand-alone PD prevalence; a more conservative patient-based definition identified PD in 191 of 334 SMAs with similar directional patterns. These outcomes are claims-based indicators of regional billing activity, not registry-equivalent estimates of modality prevalence [19].

Third, explanatory variables spanned different levels of dialysis specificity (Methods; Supplementary Table S3), so individual coefficients should be read as elements of comparative patterns across domains, modalities, and diffusion stages rather than as isolated mechanisms. The lagged HHD volume association in the PD scale model was attenuated under FDR adjustment and should be regarded as suggestive.

Finally, generalizability beyond Japan requires caution. Japan’s national insurance and reimbursement framework provides a useful setting for examining regional variation under shared payment rules, but dialysis organization, home-care infrastructure, hospital pathways, and reimbursement systems differ across institutional arrangements, and the specific associations observed here may not apply directly to other health systems.

Despite these limitations, the regional ecological design was appropriate for comparing service-system patterns across modalities and diffusion stages, and the findings identify system-level patterns that can guide longitudinal, facility-level, and patient-level research on how complex home-based care diffuses across regional health systems.

## Conclusion

Home dialysis diffusion in Japan was characterized by modality- and stage-specific service-system conditions, rather than a single regional pattern shared by PD and HHD. PD was linked mainly to existing dialysis-service organization, whereas HHD was linked to broader regional supports for sustaining more technically demanding treatment in the home. Conditions related to regional presence also differed from those related to post-presence scale, suggesting that expansion of home dialysis may require attention to the specific service-system functions needed for each modality and stage of diffusion.

## Abbreviations

CE: clinical engineer
CI: confidence interval
DID: densely inhabited district
FY: fiscal year
HD: hemodialysis
HHD: home hemodialysis
HVN: home-visit nursing agency
IRR: incidence rate ratio
LISA: local indicators of spatial association
LTC: long-term care
NDB: National Database of Health Insurance Claims and Specific Health Checkups of Japan
OR: odds ratio
PD: peritoneal dialysis
SMA: Secondary Medical Area

## Data Availability

All data referred to in this manuscript were derived from publicly available aggregate datasets. No individual-level or restricted-access data were used. Detailed source information and links to the original public data sources are provided in the Supplementary Materials.

https://www.mhlw.go.jp/stf/seisakunitsuite/bunya/0000177221_00016.html

https://www.e-stat.go.jp/

https://nlftp.mlit.go.jp/

https://kouseikyoku.mhlw.go.jp/

https://www.mhlw.go.jp/stf/kaigo-kouhyou_opendata.html

https://www.mhlw.go.jp/stf/seisakunitsuite/bunya/open_data_00016.html

## ACKNOWLEDGMENTS

The author thanks Prof. Y. Sakumura of Nara Institute of Science and Technology and Prof. K. Naemura of Tokyo University of Engineering for their general academic encouragement.

## Additional Information and Declarations

### Ethics Approval and Consent to Participate

This study used only anonymized, publicly available aggregate data. Institutional review board approval and informed consent were not required.

## Author Contributions

The author was responsible for conceptualization, methodology, funding acquisition, data curation, formal analysis, visualization, and writing of the manuscript.

## Data Availability

All data used in this study were derived from publicly available sources. Dataset links and detailed source information are provided in the Supplementary Materials.

## REFERENCES

1. Kuluski K, Ho JW, Hans PK, Nelson ML. Community Care for People with Complex Care Needs: Bridging the Gap between Health and Social Care. Int J Integr Care. 2017;17:2. 10.5334/ijic.2944.

2. Savitz LA, Bayliss EA. Emerging models of care for individuals with multiple chronic conditions. Heal Serv Res. 2021;56:980–9. 10.1111/1475-6773.13774.

3. Rose L, McKim DA, Katz SL, Leasa D, Nonoyama M, Pedersen C, et al. Home Mechanical Ventilation in Canada: A National Survey. Respir Care. 2015;60:695–704. 10.4187/respcare.03609.

4. Agar JW, Perkins A, Heaf JG. Home hemodialysis: Infrastructure, water, and machines in the home. Hemodial Int. 2015;19:S93–111. 10.1111/hdi.12290.

5. Norris AH, Shrestha NK, Allison GM, Keller SC, Bhavan KP, Zurlo JJ, et al. 2018 Infectious Diseases Society of America Clinical Practice Guideline for the Management of Outpatient Parenteral Antimicrobial Therapya. Clin Infect Dis. 2019;68:e1–35. 10.1093/cid/ciy745.

6. Mohammed SA, Roberts N, Nicolás D, Unwin S, Cotta M, Roberts JA, et al. Implementation of outpatient parenteral antimicrobial therapy program in the contemporary health care system: A narrative review of the evidence. J Infect Public Heal. 2025;18:102938. 10.1016/j.jiph.2025.102938.

7. Perl J, Brown EA, Chan CT, Couchoud C, Davies SJ, Kazancioğlu R, et al. Home dialysis: conclusions from a Kidney Disease: Improving Global Outcomes (KDIGO) Controversies Conference. Kidney Int. 2023;103:842–58. 10.1016/j.kint.2023.01.006.

8. Greenhalgh T, Robert G, Macfarlane F, BATE P, KYRIAKIDOU O. Diffusion of Innovations in Service Organizations: Systematic Review and Recommendations. Milbank Q. 2004;82:581–629. 10.1111/j.0887-378x.2004.00325.x.

9. Damschroder LJ, Aron DC, Keith RE, Kirsh SR, Alexander JA, Lowery JC. Fostering implementation of health services research findings into practice: a consolidated framework for advancing implementation science. Implement Sci. 2009;4:50. 10.1186/1748-5908-4-50.

10. Proctor E, Silmere H, Raghavan R, Hovmand P, Aarons G, Bunger A, et al. Outcomes for Implementation Research: Conceptual Distinctions, Measurement Challenges, and Research Agenda. Adm Polic Ment Heal Ment Heal Serv Res. 2011;38:65–76. 10.1007/s10488-010-0319-7.

11. Figueiredo AE, Bernardini J, Bowes E, Hiramatsu M, Price V, Su C, et al. A Syllabus for Teaching Peritoneal Dialysis to Patients and Caregivers. Perit Dial Int. 2016;36:592–605. 10.3747/pdi.2015.00277.

12. Woodrow G, Fan SL, Reid C, Denning J, Pyrah AN. Renal Association Clinical Practice Guideline on peritoneal dialysis in adults and children. BMC Nephrol. 2017;18:333. 10.1186/s12882-017-0687-2.

13. Bieber SD, Young BA. Home Hemodialysis: Core Curriculum 2021. Am J Kidney Dis. 2021;78:876–85. 10.1053/j.ajkd.2021.01.025.

14. Abra GE, Weinhandl ED, Hussein WF. Setting Up Home Dialysis Programs. Clin J Am Soc Nephrol. 2023;18:1490–6. 10.2215/cjn.0000000000000284.

15. Young BA, Chan C, Blagg C, Lockridge R, Golper T, Finkelstein F, et al. How to Overcome Barriers and Establish a Successful Home HD Program. Clin J Am Soc Nephrol. 2012;7:2023–32. 10.2215/cjn.07080712.

16. Kendzia D, Lima F, Zawierucha J, Busink E, Apel C, Malyszko JS, et al. Main Barriers to the Introduction of a Home Haemodialysis Programme in Poland: A Review of the Challenges for Implementation and Criteria for a Successful Programme. J Clin Med. 2022;11:4166. 10.3390/jcm11144166.

17. Quinn RR, Lam NN. Home Dialysis in North America. Clin J Am Soc Nephrol. 2023;18:1351–8. 10.2215/cjn.0000000000000273.

18. Johansen KL, Chertow GM, Gilbertson DT, Ishani A, Israni A, Ku E, et al. US renal data system 2022 annual data report: epidemiology of kidney disease in the United States. Am J Kidney Dis. 2023;81:A8–11. 10.1053/j.ajkd.2022.12.001.

19. Hanafusa N, Abe M, Joki N, Ogawa T, Kanda E, Kikuchi K, et al. Annual dialysis data report 2019, JSDT renal data registry. Ren Replace Ther. 2023;9:47. 10.1186/s41100-023-00478-z.

20. Slon-Roblero MF, Stel VS, Sanchez-Alvarez E, Escola JM, Dias BJF, Auñón AS, et al. Trends in home dialysis over the last decade in Europe: an ERA Registry study. Nephrol Dial Transplant. 2025;41:457–75. 10.1093/ndt/gfaf171.

21. Sood MM, Tangri N, Hiebert B, Kappel J, Dart A, Levin A, et al. Geographic and facility-level variation in the use of peritoneal dialysis in Canada: a cohort study. CMAJ Open. 2014;2:E36–44. 10.9778/cmajo.20130050.

22. Damery S, Lambie M, Williams I, Coyle D, Fotheringham J, Solis-Trapala I, et al. Centre variation in home dialysis uptake: A survey of kidney centre practice in relation to home dialysis organisation and delivery in England. Perit Dial Int. 2024;44:265–74. 10.1177/08968608241232200.

23. Potts J, Pearse CM, Lambie M, Fotheringham J, Hill H, Coyle D, et al. Patient and Center Factors in Home Dialysis Therapy Uptake: Analysis of a UK Renal Registry Cohort and a National Dialysis Center Survey. Am J Kidney Dis. 2026;87:53–64.e1. 10.1053/j.ajkd.2025.08.012.

24. Medicine C on GAF in MPB on HCSI of. Geographic Adjustment in Medicare Payment: Phase I: Improving Accuracy, Second Edition. National Academies Press (US). 2012. 10.17226/13138.

25. Ikegami N. Japan: achieving UHC by regulating payment. Glob Heal. 2019;15 Suppl 1:72. 10.1186/s12992-019-0524-4.

26. Naylor CD, Boozary A, Adams O. Canadian federal–provincial/territorial funding of universal health care: fraught history, uncertain future. CMAJ. 2020;192:E1408–12. 10.1503/cmaj.200143.

27. Hosokawa R, Ojima T, Myojin T, Aida J, Kondo K, Kondo N. Associations between Healthcare Resources and Healthy Life Expectancy: A Descriptive Study across Secondary Medical Areas in Japan. Int J Environ Res Public Heal. 2020;17:6301. 10.3390/ijerph17176301.

28. Ministry of Health, Labour and Welfare, Japan. Survey of Medical Institutions. https://www.e-stat.go.jp/en/statistics/00450021. Accessed 19 Apr 2026.

29. Ministry of Health, Labour and Welfare, Japan. The 10th NDB Open Data Japan: Part 1, Explanatory Notes. 2025. https://www.mhlw.go.jp/content/12400000/001492909.pdf. Accessed 17 Jun 2026.

30. Suzuki KT, Naemura K, Sakumura Y. Vascular access dysfunction incidence among Japanese dialysis patients from NDB Open Data Japan. Sci Rep. 2025;15:6523. 10.1038/s41598-025-91034-8.

31. Dormann CF, Elith J, Bacher S, Buchmann C, Carl G, Carré G, et al. Collinearity: a review of methods to deal with it and a simulation study evaluating their performance. Ecography. 2013;36:27–46. 10.1111/j.1600-0587.2012.07348.x.

32. Firth D. Bias Reduction of Maximum Likelihood Estimates. Biometrika. 1993;80:27. 10.2307/2336755.

33. Grogger JT, Carson RT. Models for truncated counts. J Appl Econ. 1991;6:225–38. 10.1002/jae.3950060302.

34. Moran PAP. Notes on continuous stochastic phenomena. Biometrika. 1950;37:17–23. 10.1093/biomet/37.1-2.17.

35. Anselin L. Local Indicators of Spatial Association—LISA. Geogr Anal. 1995;27:93–115. 10.1111/j.1538-4632.1995.tb00338.x.

36. Gelman A, Hill J, Yajima M. Why We (Usually) Don’t Have to Worry About Multiple Comparisons. J Res Educ Eff. 2012;5:189–211. 10.1080/19345747.2011.618213.

37. Benjamini Y, Hochberg Y. Controlling the False Discovery Rate: A Practical and Powerful Approach to Multiple Testing. J R Stat Soc: Ser B (Methodol). 1995;57:289–300. 10.1111/j.2517-6161.1995.tb02031.x.

38. Sakamoto H, Rahman MdM, Nomura S, Okamoto E, Koike S, Yasunaga H, et al. Front Matter. World Health Organization; 2018.

39. Kim B, Li Y, Lee M, Bae S, Blum MF, Le D, et al. Neighborhood Built Environment and Home Dialysis Utilization: Varying Patterns by Urbanicity-Dependent Patterns and Implications for Policy. Am J Kidney Dis. 2025;85:737–44. 10.1053/j.ajkd.2025.01.015.

40. Reddy YNV, Kearney MD, Ward M, Burke RE, O’Hare AM, Reese PP, et al. Identifying Major Barriers to Home Dialysis (The IM-HOME Study): Findings From a National Survey of Patients, Care Partners, and Providers. Am J Kidney Dis. 2024;84:567–581.e1. 10.1053/j.ajkd.2024.04.007.

41. Chan C, Combes G, Davies S, Finkelstein F, Firanek C, Gomez R, et al. Transition between Different Renal Replacement Modalities: Gaps in Knowledge and Care—the Integrated Research Initiative. Perit Dial Int. 2018;39:4–12. 10.3747/pdi.2017.00242.

42. Imbeault B, Nadeau-Fredette A-C. Optimization of Dialysis Modality Transitions for Improved Patient Care. Can J Kidney Heal Dis. 2019;6:2054358119882664. 10.1177/2054358119882664.

43. Couchoud C, Béchade C, Kolko A, Baudoin AC, Bayer F, Rabilloud M, et al. Dialysis-network variability in home dialysis use not explained by patient characteristics: a national registry-based cohort study in France. Nephrol Dial Transplant. 2022;37:1962–73. 10.1093/ndt/gfac055.

44. Poinen K, Mitra S, Quinn RR. The integrated care model: facilitating initiation of or transition to home dialysis. Clin Kidney J. 2024;17 Supplement_1:i13–20. 10.1093/ckj/sfae076.

45. Desbiens L-C, Bargman JM, Chan CT, Nadeau-Fredette A-C. Integrated home dialysis model: facilitating home-to-home transition. Clin Kidney J. 2024;17 Supplement_1:i21–33. 10.1093/ckj/sfae079.

