## Supplementary Material for "Regional Service-System Conditions Associated with Facility-Linked Home-Based Specialist Care in Japan: A Claims-Based Ecological Study of Home Dialysis"

Kenta T. Suzuki[1,2,*]

1 Graduate School of Science and Technology, Nara Institute of Science and Technology
2 Data Science Center, Nara Institute of Science and Technology, Nara, Japan

#### Supplementary Methods

##### Identification and geospatial processing of dialysis facilities

Dialysis-providing medical institutions were identified from publicly available administrative datasets (**Supplementary Table 1**). Facilities that had submitted notifications related to dialysis services were extracted and linked to a registry of insured medical institutions based on facility name and location, and address information was assigned.

For record linkage, facility names and address strings were standardized (e.g., unification of full-width and half-width characters and removal of symbols and legal entity designations), and exact matching was used as the primary criterion. Records that did not match exactly were individually reviewed using facility names and location information, and manual linkage was performed only when they were clearly identified as the same facility. Geocoding was subsequently conducted using the assigned address information to obtain longitude and latitude coordinates for each facility. Using these coordinates, dialysis facilities were spatially joined to the 2020 medical service area boundary dataset based on the within rule and assigned to secondary medical service areas.

##### Identification and spatial assignment of home-visit nursing agencies

Information on home-visit nursing agencies was obtained from publicly available administrative datasets (**Supplementary Table 1**). The published longitude and latitude data were used, and for agencies with missing coordinate information, geocoding based on address data was performed to supplement the missing values.

Home-visit nursing agencies were assigned to secondary medical service areas in the same manner as dialysis facilities, by performing a spatial join with the medical service area boundary dataset.

##### Estimation of hemodialysis volume

The volume of facility-based hemodialysis (HD) was estimated using a previously published formula. In the original method, the numbers of outpatient and inpatient HD patients were estimated as follows:

$$ESRD_{Out} = \frac{M + 2V}{12} \quad (1)$$

$$ESRD_{In} = \frac{A}{(365/7) \times 3} \quad (2)$$

$$ESRD = ESRD_{Out} + ESRD_{In} \quad (3)$$

where $ESRD_{Out}$ denotes the estimated number of outpatient HD patients, $ESRD_{In}$ the estimated number of inpatient HD patients, $M$ the number of claims for the outpatient management fee for chronic maintenance dialysis, $V$ the number of vascular access creation procedures at dialysis initiation, and $A$ the number of inpatient dialysis claims.

In the original method, the outpatient management fee for chronic maintenance dialysis cannot be claimed during the first two months after dialysis initiation under the reimbursement system. Consequently, newly initiated patients are not captured in $M$. To compensate for this structural undercount, the original formula incorporated the number of vascular access creation procedures and applied the adjustment term $M + 2V$.

In the present study, the publicly available 10th National Database (NDB) Open Data provide the cumulative number of unique patients who were charged the outpatient management fee for chronic maintenance dialysis. Therefore, $M$ in the original formula was replaced with this published unique patient count. Because this indicator directly includes patients in the early phase after dialysis initiation, the additional adjustment using $2V$ was considered unnecessary. The estimation of inpatient HD patients ($ESRD_{In}$) followed the original method.

Owing to the structure of claims-based data, the estimated values may include patients receiving home hemodialysis. In addition, the estimates may be affected by discrepancies between claims-based counts and the actual number of patients, as well as by uncertainty in the estimation of inpatient HD patients. Accordingly, the estimated HD volume was used primarily to assess relative regional differences rather than to determine absolute patient numbers.

### Supplementary Figures

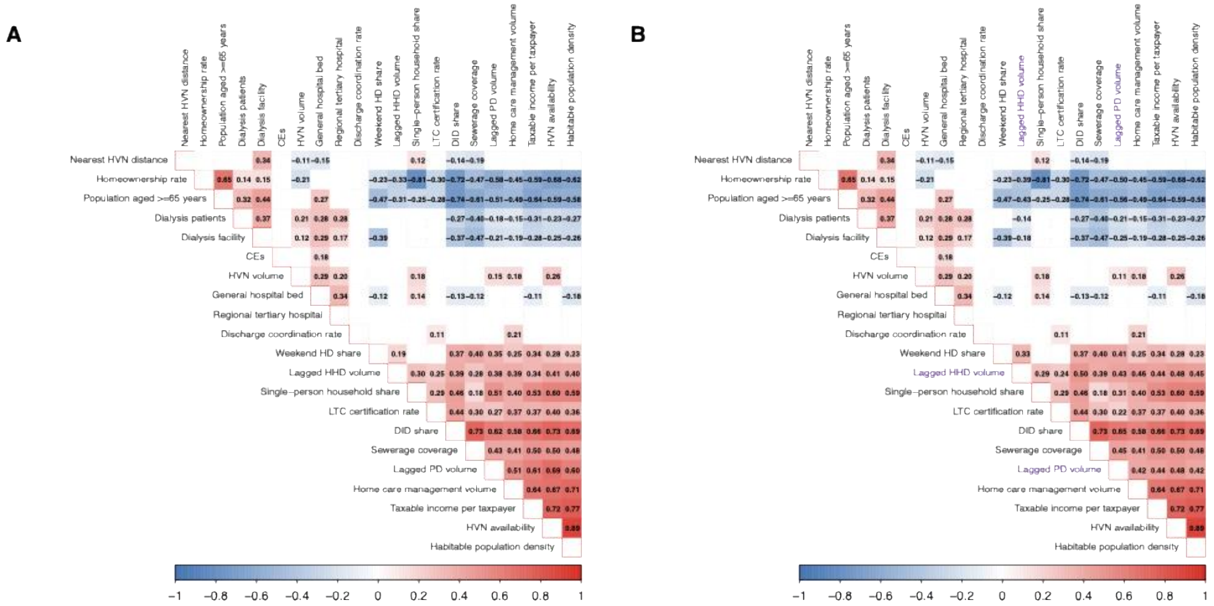


**Supplementary Figure 1 Pearson correlation matrix of candidate explanatory variables.**
This heatmap shows the Pearson correlation coefficients among the 21 candidate explanatory variables across secondary medical areas (n = 334). Panel A shows correlations using the original lagged PD and HHD claim counts, and Panel B shows correlations after log transformation of the lagged PD and HHD claim counts. The lagged home-dialysis volume variables are highlighted in purple. The variables capture dialysis care characteristics, regional healthcare and long-term care infrastructure, home-care coordination functions, and sociodemographic context. The color scale represents the direction and magnitude of correlations, ranging from −1.00 (blue) to 1.00 (red). Overall, 22 variable pairs showed strong correlations (|r| > 0.7), indicating potential multicollinearity and supporting the prespecified covariate screening procedures described in the Methods. Detailed definitions, data sources, and calculation methods for all variables are provided in **Supplementary Table 3**.

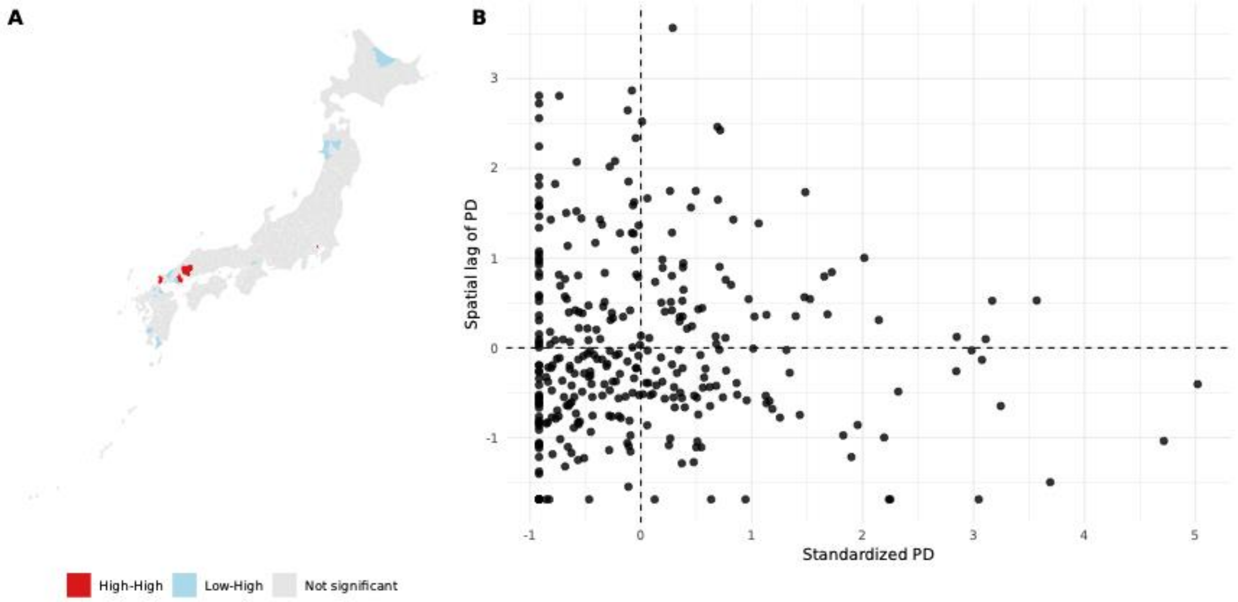


**Supplementary Figure 2 Local Indicators of Spatial Association (LISA) for PD utilization.**
Panel A shows LISA cluster maps, and Panel B shows Moran scatter plots. Colors indicate statistically significant clusters (High–High and Low–Low) at the 5% level. Most areas were not statistically significant.

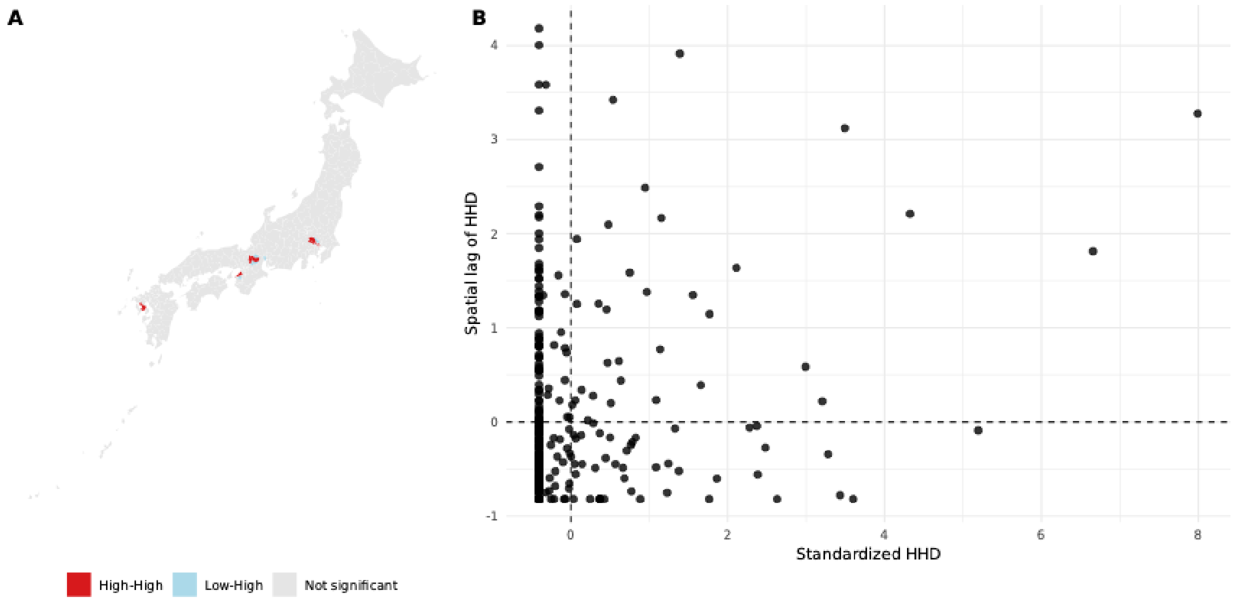


**Supplementary Figure 3 Local Indicators of Spatial Association (LISA) for HHD utilization.**
Panel A shows the LISA cluster map, and Panel B shows the Moran scatter plot. Colors indicate statistically significant clusters at the 5% level. A small number of localized clusters were observed, while most areas were not statistically significant.

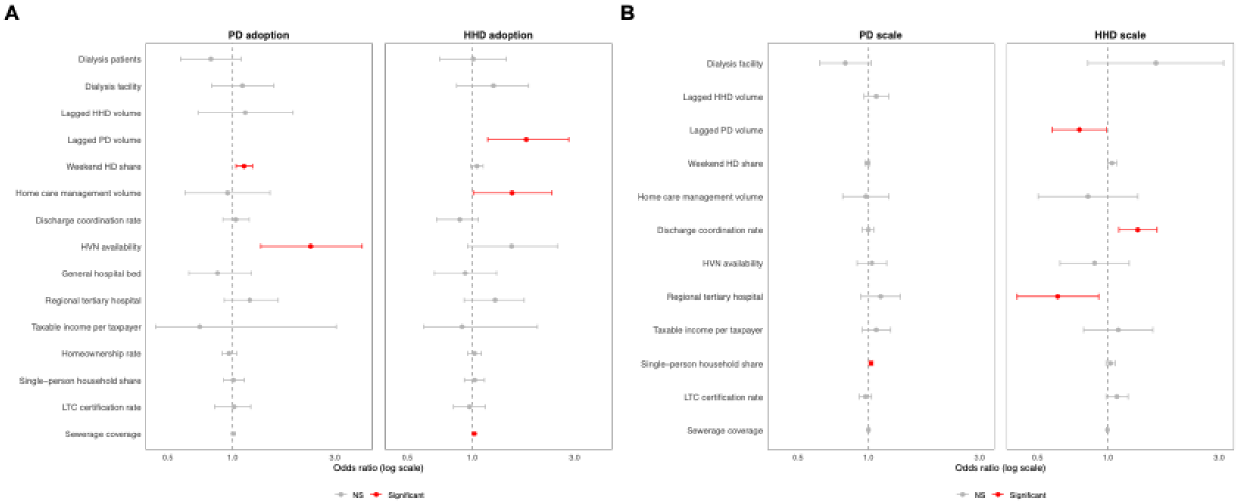


**Supplementary Figure 4 Sensitivity analyses using a 3-km threshold for proximity-based indicators.** Panel A shows adoption models, and Panel B shows scale models. Results are shown for PD adoption, HHD adoption, PD scale, and HHD scale after recalculating proximity-based indicators using a 3-km threshold. Points indicate adjusted odds ratios for adoption models and adjusted incidence rate ratios for scale models; horizontal lines indicate 95% confidence intervals. All other model specifications were unchanged from the main analysis. The main directional patterns were generally similar to those of the main analysis. Residual spatial autocorrelation was not evident for the HHD adoption model, whereas some residual spatial structure remained for the PD adoption model under this specification: response residual Moran's I = 0.076, Monte Carlo p = 0.054; Pearson residual Moran's I = 0.120, Monte Carlo p = 0.014.

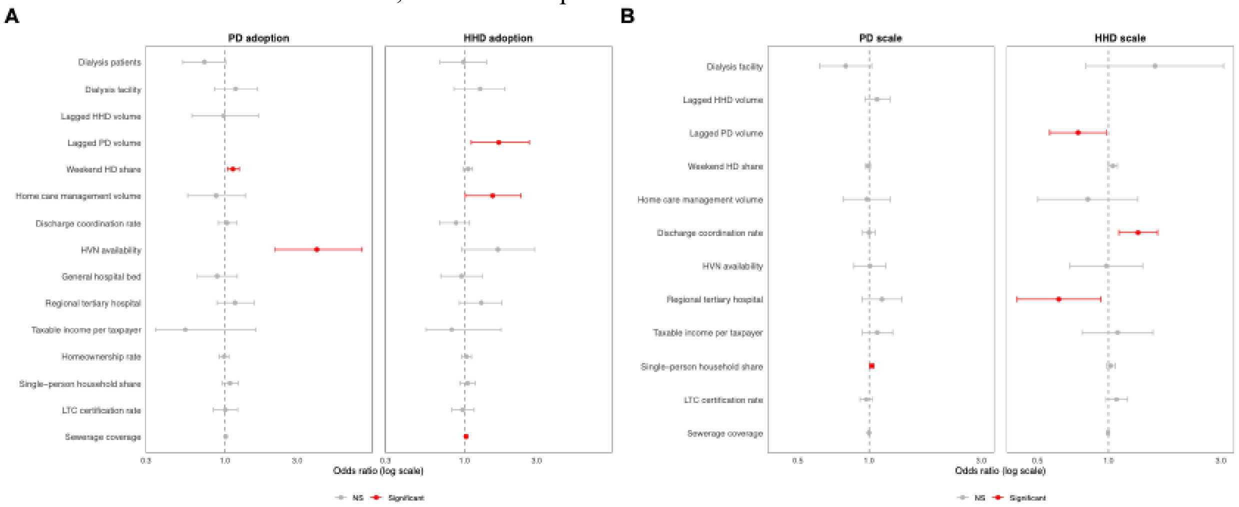


**Supplementary Figure 5 Sensitivity analyses using a 10-km threshold for proximity-based indicators.** Panel A shows adoption models, and Panel B shows scale models. Results are shown for PD adoption, HHD adoption, PD scale, and HHD scale after recalculating proximity-based indicators using a 10-km threshold. Points indicate adjusted odds ratios for adoption models and adjusted incidence rate ratios for scale models; horizontal lines indicate 95% confidence intervals. All other model specifications were unchanged from the main analysis. The main directional patterns were generally similar to those of the main analysis, although several estimates were attenuated. For the PD adoption model, some residual spatial structure remained under this specification: response residual Moran's I = 0.105, Monte Carlo p = 0.021; Pearson residual Moran's I = 0.149, Monte Carlo p = 0.014. Residual spatial autocorrelation was not evident for the PD scale model, for example, response residual Moran's I = 0.023, Monte Carlo p = 0.325; Pearson residual Moran's I = −0.030, Monte Carlo p = 0.689. Spatial diagnostics for the HHD scale model should be interpreted cautiously because the contiguity structure among adopting SMAs was sparse.

### Supplementary Tables

**Supplementary Table 1 Data items, aggregation units, and data sources**

| Item | Unit | Year | Source |
|---|---|---|---|
| **Geographic unit** | | | |
| SMA boundary / crosswalk | SMA | 2020 | [1], [2] |
| **Claims and utilization items** | | | |
| Artificial kidney | SMA | FY2023 | [3] |
| After-hours/holiday add-on (Artificial kidney) | SMA | FY2023 | [3] |
| Outpatient medical management fee for chronic maintenance dialysis patients | SMA | FY2023 | [3] |
| Home peritoneal dialysis self-care instruction/management fee | SMA | FY2022<br>FY2023 | [3] |
| Home hemodialysis instruction/management fee | SMA | FY2022<br>FY2023 | [3] |
| Comprehensive home medical management fee | SMA | FY2023 | [3] |
| Discharge joint guidance fee (Type 2) | SMA | FY2023 | [3] |
| Estimated number of discharged patients from hospitals (by patient residence), by SMA × major disease category | SMA | FY2023 | [4] |
| **Dialysis service delivery system** | | | |
| List of insured medical institutions | Facility-level | 2025 | [6] |
| Status of accepted notifications for facility standards (overall) | Facility-level | 2025 | [7] |
| Number of clinical engineers | Hospital-level | 2023 | [9] |
| **Implementation support for home-based care** | | | |
| List of home visiting-nurse stations | Facility-level | 2025 | [8] |
| Number of home-care support hospitals | Municipality | 2023 | [7] |
| Number of home-care support clinics | Municipality | 2023 | [7] |
| Home-care support clinics: number of registered facilities | Municipality | 2023 | [5] |
| Home-care support clinics: number of partner medical institutions, etc. | Municipality | 2023 | [5] |
| Home-care support clinics: number of home-care patients covered | Municipality | 2023 | [5] |
| **Hospital backup capacity** | | | |
| Number of general hospital beds | Municipality | 2024 | [5] |
| Number of regional medical support hospitals | Municipality | 2024 | [5] |
| Number of general clinics | Municipality | 2024 | [5] |
| **Living and sociodemographic context** | | | |
| Total population | Municipality | 2024 | [12] |
| Labor force population | Municipality | 2020 | [11] |
| Population aged ≥65 years | Municipality | 2020 | [11] |
| Number of households | Municipality | 2020 | [11] |
| Homeownership | Municipality | 2020 | [11] |
| Number of taxpayers | Municipality | 2024 | [13] |
| Taxable income | Municipality | 2024 | [13] |
| Population aged 15–64 years | Municipality | 2020 | [11] |
| DID population | Municipality | 2020 | [11] |
| DID area | Municipality | 2020 | [11] |
| Long-term care insurance certification rate (support levels 1–2, age- adjusted) | Municipality | 2024 | [10] |
| Sewerage service population coverage | Municipality | 2024 | [14] |
| **Geographic context** | | | |
| Habitable area | Municipality | 2023 | [15] |

Notes: Data were aggregated at the SMA, municipality, facility, or hospital level as indicated. The specific reference year for each data item is shown in the Year column. Citation numbers in the Source column correspond to the references listed in the Supplementary Information. Sewerage service population coverage was derived from the municipal-level population coverage rates for wastewater treatment and sewerage systems at the end of fiscal year 2024. Habitable area was derived from the Statistical Observations of Municipalities and corresponds to the reported inhabitable area measure for 2023.
Abbreviations: DID, densely inhabited district; SMA, secondary medical area.

**Supplementary Table 2 Detailed definitions of hemodialysis and related management fee items**

| Fee item | Claim code | No. of claims | No. of patients | Remarks (who/when the code is billed) |
|---|---|---|---|---|
| **Hemodialysis** | | | | |
| Artificial kidney | J038 | ✓ | Not available | Billed when hemodialysis is provided under the "Artificial kidney" item. Claims for the long-session add-on (other) were excluded. |
| After-hours/holiday add-on (Artificial kidney) | J038 add-on | ✓ | Not available | Add-on billed in conjunction with J038 when hemodialysis is provided under after-hours or holiday conditions. |
| Outpatient medical management fee for chronic maintenance dialysis patients | B001-2-10 | | ✓ | Billed for outpatient chronic maintenance dialysis patients when outpatient medical management is provided (patient-based). |
| **Peritoneal dialysis** | | | | |
| Home peritoneal dialysis self-care instruction/management fee | C102 | ✓ | ✓ | Billed for patients performing PD at home when self-care instruction and management are provided; includes CAPD and APD. |
| **Home hemodialysis** | | | | |
| Home hemodialysis instruction/management fee | C102-2 | ✓ | ✓ | Billed for patients receiving HHD when self-care instruction and management are provided. |
| **Home-care system** | | | | |
| Comprehensive home medical management fee | C002 | | ✓ | Billed for patients receiving home care when comprehensive medical management is provided at home; distinguished from the comprehensive management fee for residents of facilities (C002-2). |
| **Discharge support / coordination** | | | | |
| Discharge joint guidance fee (Type 2) | B005 | | ✓ | Billed for inpatients at discharge when joint guidance is provided with post-discharge providers and information is shared with relevant parties; only Type 2 claims were included in this study. |

Notes. A check mark (✓) indicates that the item was included in the corresponding aggregate. "No. of patients" represents the annual unduplicated count of patients with at least one claim for the item within the fiscal year. "Not available" indicates that the patient count was not publicly available (or not accessible) for that item. The remarks provide a concise operational description for aggregation purposes; exact billing eligibility and implementation details follow the source data specifications. Abbreviations: APD, automated peritoneal dialysis; CAPD, continuous ambulatory peritoneal dialysis; HD, hemodialysis; HHD, home hemodialysis; PD, peritoneal dialysis.

**Supplementary Table 3 Definitions of explanatory variables**

| Variable | Unit | Definition (formula) | Specificity level |
|---|---|---|---|
| **Dialysis service delivery system** | | | |
| HD patient prevalence | Persons per 100,000 population | (No. of HD patients / Total population) x 100,000 | Dialysis-specific |
| Dialysis facility density | Facilities per 100,000 population | (No. of dialysis facilities / Total population) x 100,000 | Dialysis-specific |
| Clinical engineers | Clinical engineers per 100,000 population | (No. of clinical engineers / Total population) x 100,000 | Dialysis-specific |
| Weekend HD share | % | (No. of HD sessions billed with the after-hours/holiday add-on / Total no. of HD sessions) x 100 | Dialysis-specific |
| Lagged PD volume | Claims | No. of claims for home peritoneal dialysis self-care instruction/management fee in FY2022 | Dialysis-specific |
| Lagged HHD volume | Claims | No. of claims for home hemodialysis instruction/management fee in FY2022 | Dialysis-specific |
| **Implementation support for home-based care** | | | |
| Home-care support hospital density | Facilities per 100,000 population | (No. of home-care support hospitals / Total population) x 100,000 | Medical, non-dialysis-specific |
| Home care management volume | Persons per 100,000 population | (No. of persons billed for comprehensive home medical management fee / Total population) x 100,000 | Medical, non-dialysis-specific |
| Discharge coordination rate | % | (No. of patients billed for discharge joint guidance fee (Type 2) / Estimated no. of discharged patients) x 100 | Medical, non-dialysis-specific |
| Nearest HVN agency distance | km | Median straight-line distance from each dialysis facility to the nearest home-visit nursing agency within the SMA | Medical, non-dialysis-specific |
| HVN availability | Facilities | Median number of home-visit nursing agencies within a 5-km radius of each dialysis facility within the SMA | Medical, non-dialysis-specific |
| **Hospital backup capacity** | | | |
| General hospital bed density | Beds per 100,000 population | (No. of general hospital beds / Total population) x 100,000 | Medical, non-dialysis-specific |
| Regional tertiary hospital density | Facilities per 100,000 population | (No. of regional medical support hospitals / Total population) x 100,000 | Medical, non-dialysis-specific |
| **Living and sociodemographic context** | | | |
| Habitable population density | Persons per km2 of habitable area | Total population / Habitable area | General regional context |
| DID share | % | (Population in DID / Total population) x 100 | General regional context |
| Taxable income per taxpayer | JPY 1,000 per person | Taxable income / No. of taxpayers, expressed in units of JPY 1,000 | General regional context |
| Homeownership rate | % | (No. of owner-occupied households / No. of households) x 100 | General regional context |
| Single-person household share | % | (No. of single-person households / No. of households) x 100 | General regional context |
| Population aged ≥65 years | % | (Population aged ≥65 years / Total population) x 100 | General regional context |
| LTC certification rate | % | Age-adjusted percentage of persons certified as needing long-term care/support, as reported in official statistics | General regional context |
| Sewerage coverage | % | (Population covered by sewerage service / Total population) x 100 | General regional context |

Notes. Older adults were defined as persons aged ≥65 years. Percentage variables were calculated as proportions multiplied by 100. The holiday add-on is specific to HD (hemodialysis) in the study setting; therefore, the holiday add-on rate was defined using HD sessions. Distance-based measures used straight-line distance; the reference location and the aggregation unit should be specified to ensure reproducibility. Labor force population (aged ≥15 years) should be operationally defined according to the data source (e.g., employed plus unemployed actively seeking work), because definitions vary across statistical systems. The discharge joint guidance claim rate was constructed using Type 2 claims only.

Specificity level classifies each covariate by how directly it captures dialysis service delivery, based on the data source of the variable. Dialysis-specific indicators are constructed from dialysis-related claims, dialysis facilities, or dialysis-related staff. Medical, non-dialysis-specific indicators are constructed from medical claims or medical facilities that include dialysis but are not limited to it. General regional context indicators are constructed from non-medical sources, including population, housing, taxation, and infrastructure data. The Note column flags variables with measurement characteristics (proximity-based indicators that overlap with urban service density) or conceptual relevance to home dialysis requirements (sewerage coverage, single-person household share) that warrant additional interpretive caution.

Abbreviations: DID, densely inhabited district; HD, hemodialysis; HHD, home hemodialysis; LTC, long-term care; PD, peritoneal dialysis; HVN, home-visit nursing agency.

**Supplementary Table 4 Firth penalized logistic regression for PD adoption.**

| Variable | Adjusted OR | 95% CI | P value | q value |
|---|---|---|---|---|
| Dialysis patients | 0.76 | 0.55–1.05 | 0.096 | 0.447 |
| Dialysis facility | 1.13 | 0.82–1.57 | 0.467 | 0.755 |
| Lagged HHD volume | 1.08 | 0.68–1.81 | 0.762 | 0.820 |
| Weekend HD share | 1.13 | 1.04–1.24 | 0.004 | 0.026 |
| Home care management volume | 0.91 | 0.59–1.43 | 0.688 | 0.803 |
| Discharge coordination rate | 1.04 | 0.91–1.21 | 0.539 | 0.755 |
| HVN availability | 2.75 | 1.57–4.96 | <0.001 | 0.006 |
| General hospital bed | 0.88 | 0.65–1.23 | 0.395 | 0.755 |
| Regional tertiary hospital | 1.17 | 0.89–1.58 | 0.268 | 0.755 |
| Taxable income per taxpayer | 0.65 | 0.41–2.51 | 0.535 | 0.755 |
| Homeownership rate | 0.97 | 0.90–1.05 | 0.388 | 0.755 |
| Single-person household share | 1.02 | 0.92–1.15 | 0.667 | 0.803 |
| LTC certification rate | 1.00 | 0.82–1.20 | 0.991 | 0.991 |
| Sewerage coverage | 1.01 | 0.99–1.02 | 0.356 | 0.755 |

Model fit: likelihood ratio test, $\chi^2 = 71.81$, df = 14, $p < 0.001$; n = 334.

**Supplementary Table 5 Firth penalized logistic regression for HHD adoption.**

| Variable | Adjusted OR | 95% CI | P value | q value |
|---|---|---|---|---|
| Dialysis patients | 0.99 | 0.69–1.41 | 0.961 | 0.961 |
| Dialysis facility | 1.25 | 0.84–1.82 | 0.261 | 0.457 |
| Lagged PD volume | 1.73 | 1.14–2.74 | 0.009 | 0.081 |
| Weekend HD share | 1.05 | 0.98–1.13 | 0.142 | 0.332 |
| Home care management volume | 1.56 | 1.03–2.37 | 0.036 | 0.170 |
| Discharge coordination rate | 0.88 | 0.69–1.07 | 0.234 | 0.457 |
| HVN availability | 1.53 | 0.92–2.59 | 0.101 | 0.332 |
| General hospital bed | 0.94 | 0.68–1.31 | 0.727 | 0.794 |
| Regional tertiary hospital | 1.28 | 0.92–1.74 | 0.141 | 0.332 |
| Taxable income per taxpayer | 0.87 | 0.58–1.93 | 0.656 | 0.794 |
| Homeownership rate | 1.02 | 0.95–1.10 | 0.520 | 0.794 |
| Single-person household share | 1.03 | 0.92–1.15 | 0.607 | 0.794 |
| LTC certification rate | 0.97 | 0.82–1.16 | 0.738 | 0.794 |
| Sewerage coverage | 1.02 | 1.00–1.04 | 0.012 | 0.081 |

Model fit: likelihood ratio test, $\chi^2 = 98.91$, df = 14, $p < 0.001$; n = 334.

**Supplementary Table 6 Zero-truncated negative binomial regression for PD scale.**

| Variable | Adjusted IRR | 95% CI | P value | q value |
|---|---|---|---|---|
| Dialysis facility | 0.67 | 0.53–0.84 | <0.001 | 0.004 |
| Lagged HHD volume | 1.14 | 1.01–1.29 | 0.030 | 0.122 |
| Weekend HD share | 0.99 | 0.97–1.02 | 0.598 | 0.740 |
| Home-care support hospital | 1.02 | 0.86–1.22 | 0.793 | 0.843 |
| Home care management volume | 0.96 | 0.83–1.11 | 0.599 | 0.740 |
| Discharge coordination rate | 1.02 | 0.95–1.08 | 0.617 | 0.740 |
| HVN availability | 1.12 | 0.97–1.30 | 0.148 | 0.444 |
| Regional tertiary hospital | 1.12 | 0.94–1.33 | 0.200 | 0.462 |
| Taxable income per taxpayer | 1.09 | 0.94–1.24 | 0.231 | 0.462 |
| Single-person household share | 1.03 | 1.01–1.05 | 0.002 | 0.009 |
| LTC certification rate | 0.98 | 0.92–1.04 | 0.407 | 0.699 |
| Sewerage coverage | 1.00 | 0.99–1.00 | 0.843 | 0.843 |

Dispersion parameter: $\theta = 1.67$; Log-likelihood: −1795. Values are adjusted incidence rate ratios from a zero-truncated negative binomial regression model for PD scale among adopting SMAs. The logarithm of the total number of dialysis patients was included as an offset. q values were calculated using the Benjamini–Hochberg false discovery rate procedure within this model.

**Supplementary Table 7 Zero-truncated negative binomial regression for HHD scale.**

| Variable | Adjusted IRR | 95% CI | P value | q value |
|---|---|---|---|---|
| Dialysis facility | 1.12 | 0.70–1.80 | 0.640 | 0.640 |
| Lagged PD volume | 0.86 | 0.65–1.14 | 0.296 | 0.510 |
| Weekend HD share | 1.03 | 0.99–1.08 | 0.131 | 0.393 |
| Home-care support hospital | 1.15 | 0.86–1.54 | 0.338 | 0.510 |
| Home care management volume | 0.88 | 0.62–1.25 | 0.489 | 0.586 |
| Discharge coordination rate | 1.33 | 1.11–1.60 | 0.002 | 0.029 |
| HVN availability | 0.84 | 0.60–1.19 | 0.340 | 0.510 |
| Regional tertiary hospital | 0.64 | 0.47–0.87 | 0.005 | 0.029 |
| Taxable income per taxpayer | 1.09 | 0.78–1.52 | 0.604 | 0.640 |
| Single-person household share | 1.03 | 0.98–1.08 | 0.227 | 0.510 |
| LTC certification rate | 1.09 | 0.98–1.21 | 0.095 | 0.379 |
| Sewerage coverage | 1.00 | 0.98–1.01 | 0.385 | 0.513 |

Dispersion parameter: $\theta = 1.30$; Log-likelihood: −562.9. Values are adjusted incidence rate ratios from a zero-truncated negative binomial regression model for HHD scale among adopting SMAs. The logarithm of the total number of dialysis patients was included as an offset. IRRs are presented per one-unit increase in each covariate as coded in the model. q values were calculated using the Benjamini–Hochberg false discovery rate procedure within this model.

**Supplementary Table 8 Spatial autocorrelation of model residuals assessed using Moran's I based on response residuals.**

Moran's I was calculated using response residuals from each fitted model.

| Model | Moran's I | P value | Interpretation |
|---|---|---|---|
| PD adoption | 0.056 | 0.059 | Borderline positive spatial autocorrelation |
| HHD adoption | 0.038 | 0.141 | No significant spatial autocorrelation |
| PD scale | 0.007 | 0.410 | No significant spatial autocorrelation |
| HHD scale | -0.025 | 0.490 | No significant spatial autocorrelation |

**Supplementary Table 9 Sensitivity analysis for PD adoption using patient-based presence instead of claims-based presence.**

| Variable | Adjusted OR | 95% CI | P value |
|---|---|---|---|
| Dialysis patients | 0.70 | 0.51–0.95 | 0.023 |
| Dialysis facility | 0.97 | 0.69–1.35 | 0.879 |
| Lagged HHD volume | 1.54 | 1.07–2.26 | 0.018 |
| Weekend HD share | 1.09 | 1.01–1.17 | 0.018 |
| Home care management volume | 1.15 | 0.77–1.72 | 0.501 |
| Discharge coordination rate | 1.13 | 0.99–1.29 | 0.080 |
| HVN availability | 2.26 | 1.41–3.71 | <0.001 |
| General hospital bed | 1.30 | 0.97–1.76 | 0.084 |
| Regional tertiary hospital | 1.33 | 1.00–1.79 | 0.048 |
| Taxable income per taxpayer | 1.15 | 0.44–2.79 | 0.758 |
| Homeownership rate | 0.99 | 0.93–1.06 | 0.818 |
| Single-person household share | 1.08 | 0.97–1.21 | 0.143 |
| LTC certification rate | 0.97 | 0.81–1.14 | 0.683 |
| Sewerage coverage | 1.00 | 0.99–1.02 | 0.537 |

Note: PD adoption was redefined using patient-based presence rather than claims-based presence. Specifically, SMAs were classified as adopting PD when a non-suppressed patient count for the home peritoneal dialysis self-care instruction/management fee was reported in FY2023. Under this definition, 191 of 334 SMAs were classified as adopting PD. Associations were estimated using Firth penalized logistic regression. The overall pattern of associations was broadly similar to that of the main analysis. No significant residual spatial autocorrelation was detected in this sensitivity model (response residuals: Moran's I = −0.0406, randomization p = 0.680; Pearson residuals: Moran's I = −0.0424, randomization p = 0.690).

#### Supplementary References


1. Ministry of Health, Labour and Welfare (Japan). Secondary medical areas (SMA): municipality crosswalk (dataset). e-Gov Data Portal. https://data.e-gov.go.jp/data/dataset/mhlw_20150115_0041 Accessed May 19, 2026.
2. Ministry of Land, Infrastructure, Transport and Tourism (Japan). National Land Numerical Information: Medical service areas (A38). https://nlftp.mlit.go.jp/ksj/gml/datalist/KsjTmplt-A38.html Accessed May 19, 2026.
3. Ministry of Health, Labour and Welfare (Japan). 10th NDB Open Data (National Database of Health Insurance Claims and Specific Health Checkups of Japan), FY2022 and FY2023 claims data. https://www.mhlw.go.jp/stf/seisakunitsuite/bunya/0000177221_00016.html Accessed May 19, 2026.
4. Ministry of Health, Labour and Welfare (Japan). Patient Survey (2023): secondary medical area tables - estimated hospital discharges by SMA x major disease category (Table N22; statdisp_id=0004026159). e-Stat. https://www.e-stat.go.jp/stat-search/database?statdisp_id=0004026159 Accessed May 19, 2026.
5. Ministry of Health, Labour and Welfare (Japan). Survey of Medical Institutions. e-Stat. https://www.e-stat.go.jp/stat-search/files?tstat=000001030908 Accessed May 19, 2026.
6. Regional Bureaus of Health and Welfare, Ministry of Health, Labour and Welfare (Japan). Lists of insured medical institutions and pharmacies, by regional bureau jurisdiction. Regional Bureaus of Health and Welfare portal. https://kouseikyoku.mhlw.go.jp/ Accessed May 19, 2026.
7. Regional Bureaus of Health and Welfare, Ministry of Health, Labour and Welfare (Japan). Acceptance lists for filed facility standards, by regional bureau jurisdiction. Regional Bureaus of Health and Welfare portal. https://kouseikyoku.mhlw.go.jp/ Accessed May 19, 2026.
8. Ministry of Health, Labour and Welfare (Japan). Long-term care service information disclosure system: Open Data. https://www.mhlw.go.jp/stf/kaigo-kouhyou_opendata.html Accessed May 19, 2026.
9. Ministry of Health, Labour and Welfare (Japan). Bed Function Report: Open Data, FY2023 reporting. https://www.mhlw.go.jp/stf/seisakunitsuite/bunya/open_data_00016.html Accessed May 19, 2026.
10. Ministry of Health, Labour and Welfare (Japan). Long-term Care Insurance Business Status Report. https://www.mhlw.go.jp/toukei/list/84-1.html Accessed May 19, 2026.
11. Statistics Bureau of Japan (Ministry of Internal Affairs and Communications). Population Census 2020. e-Stat. https://www.e-stat.go.jp/stat-search/files?tstat=000001136464 Accessed May 19, 2026.
12. Ministry of Internal Affairs and Communications (Japan). Survey on population, vital events, and households based on the Basic Resident Register. e-Stat. https://www.e-stat.go.jp/stat-search/files?toukei=00200241 Accessed May 19, 2026.
13. Ministry of Internal Affairs and Communications (Japan). Local Public Finance Survey: taxation status (municipalities), Table 18 (statdisp_id=0003172930). e-Stat. https://www.e-stat.go.jp/stat-search/database?statdisp_id=0003172930 Accessed May 19, 2026.
14. Ministry of Land, Infrastructure, Transport and Tourism (Japan). List of municipal population coverage rates for wastewater treatment and sewerage systems at the end of fiscal year 2024. https://www.mlit.go.jp/mizukokudo/sewerage/content/001908906.pdf Accessed May 19, 2026.
15. Statistics Bureau of Japan (Ministry of Internal Affairs and Communications). Statistical Observations of Municipalities 2023, Basic Data, file B: Natural Environment. e-Stat. https://www.e-stat.go.jp/stat-search/files Accessed May 19, 2026.
